# Estimating the Fraction of Unreported Infections in Epidemics with a Known Epicenter: an Application to COVID-19

**DOI:** 10.1101/2020.04.13.20063511

**Authors:** Ali Hortaçsu, Jiarui Liu, Timothy Schwieg

**Affiliations:** Kenneth C. Griffin Department of Economics, University of Chicago and NBER; Kenneth C. Griffin Department of Economics, University of Chicago; Becker Friedman Institute

**Author notes:** We thank the Becker Friedman Institute for financial support. We also thank Fernando Alvarez, Susan Athey, Patrick Bayer, Rana Choi, Liran Einav, Jeremy Fox, Mikhail Golosov, Austan Goolsbee, Philip Haile, Jakub Kastl, Magne Mogstad, Casey Mulligan, Derek Neal, Robert Shimer, Jose Scheinkman, Chad Syverson, Harald Uhlig, Theodore Vassilakis, and Alessandra Voena for their helpful comments.

## Abstract

We develop a simple analytical method to estimate the fraction of unreported infections in epidemics with a known epicenter and estimate the number of unreported COVID-19 infections in the US during the first half of March 2020. Our method utilizes the covariation in initial reported infections across US regions and the number of travelers to these regions from the epicenter, along with the results of a randomized testing study in Iceland. We estimate that 4-14% (1.5%-10%) of actual infections had been reported in US up to March 16, accounting for an assumed reporting lag of 8 (5) days.

## 1 Introduction

The global pandemic COVID-19 is here in the United States. The number of confirmed cases is rising rapidly, reaching 398,809 as of April 7 with 12,895 reported deaths. The coronavirus outbreak was declared a national emergency beginning March 1^1^. More than half of U.S. states have imposed various levels of lockdown measures^2^. In addition to the public health crisis, the country is certainly looking at a deep and possibly longlasting economic recession, according to Ben Bernanke and Janet Yellen in a recent Financial Times article^3^.

Given the level of severity of current conditions, we still fail to answer the most basic yet important question: How many people are actually infected with COVID-19 in the U.S. and what is the true fatality rate? Because of the shortage in testing kits, hospitals and disease control centers are only able to test the subsample of people with severe symptoms or travel history. The number of reported infections is much lower than the actual number of infections in the U.S.

These unreported infections can be unrecognized because they often experience mild or no symptoms (Nishiura et al., 2020; Andrei, 2020). If not hospitalized or quarantined, they can infect a large proportion of the population. Thus, estimating the number of unreported infections can inform policy-makers about the proper scale of virus control policies (Alvarez et al., 2020; Eichenbaum et al., 2020), and to assess the effectiveness of public health policies such as social distancing in slowing the spread of the epidemic.

Estimating the number of unreported infections may also give a more accurate measure of the true fatality rate. The current reported fatality rate, which is 3-4% according to WHO^4^, is not the true fatality rate. The true fatality rate is the proportion of those *actually infected* who die, not of those *reportedly infected*. The reported fatality rate is a biased estimate of the true rate, because there is selection bias in testing. Since many of the patients who are tested have severe symptoms, they may have a higher true fatality rate than those untested, which would make the reported fatality rate an overestimate of the true rate.

Ideally, a randomized testing experiment will give an unbiased estimate of the true rate. However, given the limited supply of testing kits and surging demand by people with symptoms, randomized testing may be infeasible, especially in the early periods of the outbreak. Therefore, it may be of great value to estimate the fraction of unreported infections with observational data at hand. With that knowledge, policy-makers will be better equipped to assess the proper level and duration of virus control policies.

In this paper we develop a simple analytical method to estimate the fraction of unreported infections for situations where the epidemic has a known epicenter. Our methodological strategy, described in Section 3, exploits the covariation between the number of initial reported infections in locations away from the epicenter, and the number of travelers from the epicenter to these locations.

To illustrate the idea, consider a time period when the epicenter is the only location with infections, and that the only way another city/country can be infected is through travelers. Also assume, as in Section 3.2, that any infected travelers can only come from the unreported infected population in the epicenter – an assumption we find reasonable (as reported infected individuals would not be allowed to travel), but are able to relax in Section 3.3. Suppose now the hypothetical situation where we know the reporting rate of infections in the epicenter (the fraction of reported infections to the true number of infections), and that we know the number of travelers from the epicenter to another city/country. Assuming travelers resemble the population of the epicenter, we can calculate the expected number of infected (but unreported) travelers entering other cities/countries. Assuming further that we know the rate of transmission of the disease, we can then calculate the expected number of infections these travelers will have generated in these locations. Comparing the expected number of infections that arise from travelers to reported cases of the infection, we can estimate the reporting rate.

What can we do in the realistic case if the reporting rate in the epicenter is unknown? In Section 3.2, we propose the following: suppose we make the assumption that the reporting rate at the epicenter and the previously uninfected city/country are the same (or a known function of each other). We can then start with a guess on the unknown rate of reporting at the epicenter, which allows us to calculate the implied reporting rate at the previously uninfected city/country, and check whether these are equal (or satisfy the known function). If not, we update our guess, and try again. In other words, we can solve for the reporting rate(s) balancing the expected number of infections from travel and the number of infected that are being reported in both locations.

While the above strategy, outlined in Section 3.2, is in principle implementable, it is crucially dependent on the assumption that reporting rates are the same across the epicenter and destination locations (or a known function of each other). Moreover, its results are very sensitive to knowing the transmission rate of the infection from travelers, as this allows us to project the number of infections in the destination city/country. Suppose now that we have access to the reporting rate of infections from another destination city/country, e.g. through universal or randomized testing, as has been done in Iceland.^5^ This allows us to estimate how infectious the travelers from the epicenter are. Assuming that this transmission rate from travelers is the same (or a known function of) as the transmission rate at the destination city/country of interest, we can then calculate the expected number of infections we would expect from travel. Intuitively, the ratio between number of travelers to two destination cities/countries from the epicenter should tell us the ratio of total infections between the two cities/countries. Randomized or universal testing at one of the destinations, Iceland in our case, will give us its number of total infections, so total infections at the other destination can be computed. This strategy is discussed in detail in Section 3.3.

We would like to be very upfront that the estimation strategies outlined above are dependent on strong assumptions and reliable data on travel patterns, and that any results are *very sensitive to these assumptions*. However, our hope is that our approach is clear in terms of its assumptions and its corresponding limitations; we hope that future research can improve upon these limitations. We have attempted to account for some of the limitations. For example, in Section 3.4, we discuss how to correct for the fact that infections are often reported with a delay, as there is a delay to the outset of symptoms that are often a prerequisite for testing for the infection, as well as a delay in laboratory testing.

Our data consists of detailed daily reported infections for all U.S. states/city/county and Iceland collected by Johns Hopkins University of Medicine Coronavirus Resource Center from January 22 to March 31, 2020; international travel data to U.S. in January and February 2020 from I-94 travels data by National Travel and Tourism Office; international travel data to Iceland by Icelandic Tourist Board in January and February 2020.

Our model generates a range of estimates that depend on the traveler data that is incorporated, the date range considered, and assumptions regarding the lags associated with reported case data. We report this range of estimates in Table 3. Across these estimates, we find that 4% to 14% of cases were reported across the U.S. up to March 16, when social distancing measures began to be applied in major metropolitan areas and travel declined significantly (Thompson et al., 2020).^6^ This suggests that for each case reported in late February/early March, between 6 to 24 cases remained unreported (after accounting for an 8 day reporting lag). Once again, our estimates are highly dependent on model assumptions, and the data that is used to inform it. We discuss how our results depend on these assumptions in some detail in Section 5.

**Table 1:** Summary Statistics of Fraction of Reported Infections by County

| Version | Min. | 1st Qu. | Median | Mean | 3rd Qu. | Max. |
| --- | --- | --- | --- | --- | --- | --- |
| China Travel Only | 0.001404 | 0.027434 | 0.037345 | 0.060896 | 0.100897 | 0.203500 |
| China and Italian Travel | 0.001254 | 0.025142 | 0.034784 | 0.055746 | 0.094906 | 0.183048 |
| China and EU Travel | 0.001397 | 0.024920 | 0.038928 | 0.051934 | 0.067098 | 0.193740 |

Summary statistics reported on the distribution of $\alpha$ for each county in the data. $\alpha$ is estimated for $T_0$ Feb 23, $T_1$ Mar 13, and a Lag of eight days. EU travel includes traveler data from Italy, Spain, UK, and Germany.

**Table 2:**
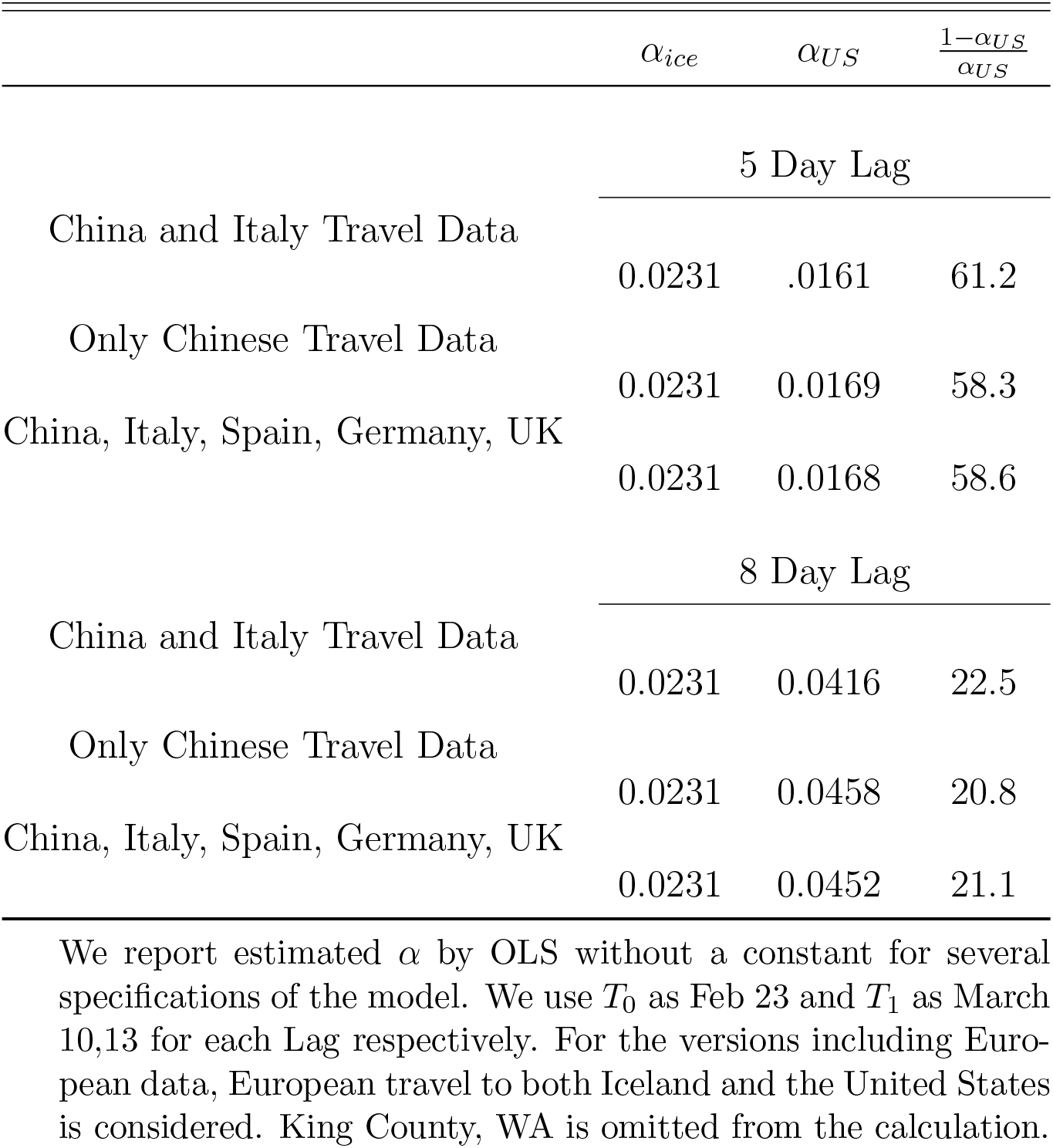
Estimated average fraction of reported infections

**Table 3:**
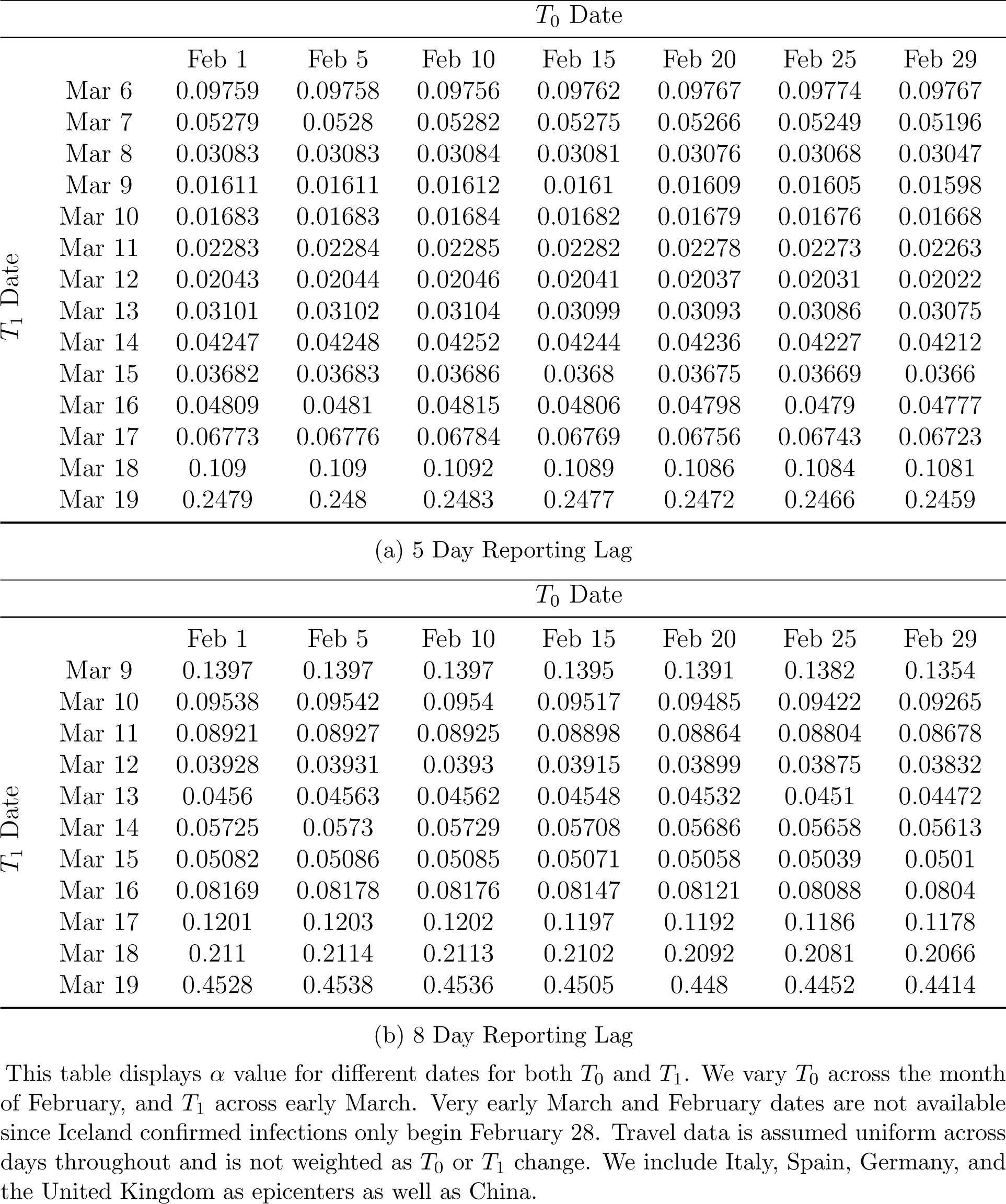
Mean Fraction of Unreported Infections with Different Cutoffs

In the economic literature, Berger et al. (2020) and Stock (2020) study the importance of unreported cases in the context of the coronavirus pandemic. Our paper contributes to the growing literature in epidemiology on estimating the true number of infections using observational data and structural model assumptions. Notably, (Li et al., 2020; Wu et al., 2020; Flaxman et al., 2020; Liu et al., 2020a,b; Nishiura et al., 2020) utilize simulated epidemiological models to estimate the fraction of unreported infections in China and European countries. As Zhao et al. (2020) notes, it is often difficult to identify the fraction of unreported alongside the growth of the infection purely by measures of fit. Our paper complements these extant papers: we provide what we believe is a transparent identification argument and a very light computational strategy that allows researchers to assess the sensitivity of model estimates to modeling and data assumptions. That said, our model may miss important components of disease dynamics that these more sophisticated epidemiological models incorporate. These richer models may also allow one to estimate a richer set of model parameters than we have been able to.^7^ Another related recent paper is Imai et al. (2020), who estimate potential total cases in Wuhan China from the confirmed cases in other countries due to international travel, assuming that all cases outside of China are reported correctly ^8^. Korolev (2020) discusses non-identification in SEIRD models and proposes estimation strategy conditional on knowing infectious period and incubation period.

Section 2 introduces our model of infection, which describes the early stages of the dynamics of the epidemic. Section 3 presents our two estimation/identification strategies. Section 4 describes the data we are using for estimation. Section 5 lays out the estimation results and our robustness checks.

## 2 Model

Our model is based on the classic SIR model in epidemiology. We consider the evolution of the virus in both the epicenter *c* and into target city *i* over a period of time *T*_0_ *≤t ≤ T*_1_. We are considering a relative short period of time in the early stage of the epidemics. Thus, the “recovered” population at the epicenter, which is a small fraction of the population, is assumed not to play a significant role during this period.

### 2.1 What happens at the epicenter *c*

We denote Infected, Reported Infected, and Unreported Infected in time *t* and epicenter *c* as *I*_*c,t*_, *R*_*c,t*_, *U*_*c,t*_ respectively.

The epicenter starts with some initial infections *I*_*c*,0_. We are considering a short period of time in between *T*_0_ and *T*_1_, so the number of susceptibles at the epicenter remain relatively constant throughout this period. There are also no infected cases traveling into epicenter. We assume no recovery. So at time *t*, the total infections at epicenter with transmission rate *β* is given by

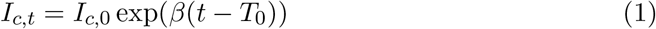

It is worth noting that this *β* term can include the spread minus recoveries, since we do not model a changing number of susceptibles. It should be viewed as the net spread of infections over time.

Each time *t*, there is a cohort of travelers *M*_*i,t*_ going from epicenter to target city and potentially bringing the virus to target city.

### 2.2 What happens in target city *i*

We denote Infected, observed Reported Infected, and Unreported Infected in time *t* and city *i* as *I*_*i,t*_, *R*_*i,t*_, *U*_*i,t*_ respectively. At period *T*_0_, target city *i* has zero infections, so 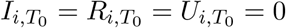.

Each time *t ∈* [*T*_0_, *T*_1_], target city receives a cohort *t* of incoming travelers *M*_*i,t*_ from the epicenter. Among these travelers, 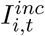 are infected. Each cohort of incoming infected 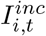 will transmit the virus in target city with rate *β* for the period of [*t, T*_1_]. We assume that the transmission rate at target city is the same as in epicenter. Thus, at period *T*_1_, this cohort will infect 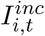 exp(*β*(*T*_1_ − *t*)) people in the city *i*. The total new infections at target city at *T*_1_ caused by all cohorts of incoming infected travelers will be

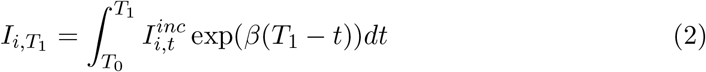

Let *α* be fraction of reported case over new infections across periods, so 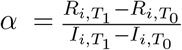. Since 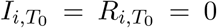, we can write 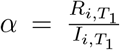. In reality at target city *i*, we only observe *R*_*i,t*_ with some iid measurement error *ϵ*_*i,t*_^9^. Let 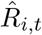 denote the observed reported cases of city *i* time *t*. We have

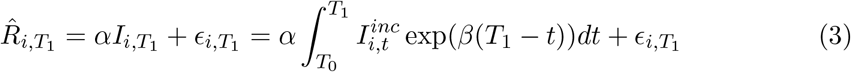

## 3 Estimating the Reporting Rate *α*

The estimation/identification question is: can we recover *α*, the reporting rate, when we only observe reported infections 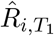 but not 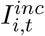, the total incoming infected in equation (3)? In the following Sections 3.1-3.3, we provide a complete treatment of how one can recover *α* under different scenarios of data availability. We consider two sets of data that could potentially be available: (i) data on travel from epicenter to U.S., and (ii) data from a randomized testing implemented outside of U.S. In Section 3.4, we extend our model and estimation strategy to incorporate reporting lags.

### 3.1 Travel data unavailable but randomized testing data available

If randomized testing data from some other country is available, the true infection rate in that country can be estimated. Therefore, given the population and number of reported infections, the fraction of reported infections *in that country* can be estimated. If we are willing to believe that the fraction of reported infections in that country is the same as in the U.S. due to similar testing availability or medical systems, then *α* is trivially recovered. However, in many cases, this assumption is unlikely to hold. In the next sections, we will show how we can recover *α* relaxing this assumption.

### 3.2 Travel data available but randomized testing data unavailable

When only travel data is available, we need the assumption that people capable of traveling away from the epicenter would be the uninfected and the unreported infected. This is a reasonable assumption especially in the case of COVID-19 because the great majority of reported infected individuals would be quarantined and not allowed to travel.

Our main assumption in this scenario is:

#### Assumption 3.1.

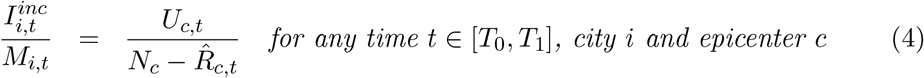

*N*_*c*_ is the population of epicenter. 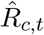 is the observed reported infections at epicenter *c* time *t*, defined analogously to 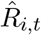 for target city *i*. In other words, we are assuming that the fraction of unreported infections among incoming travelers from the epicenter is the same as the fraction of unreported infections among people capable of leaving the epicenter. (We will relax this assumption in Section 3.3.) Since *α* remains constant,^10^ we have 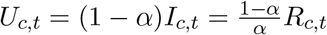. Therefore, assumption 3.1 becomes:

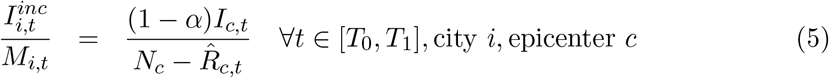

Plugging equation (1) in, we get

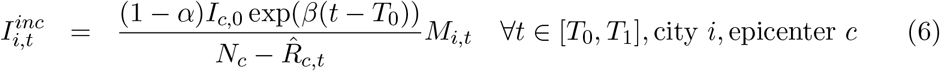

Plugging back to equation (3), we get

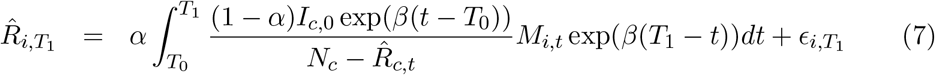

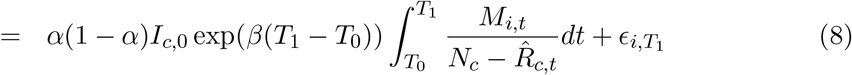

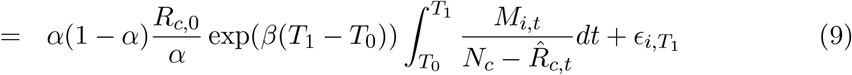

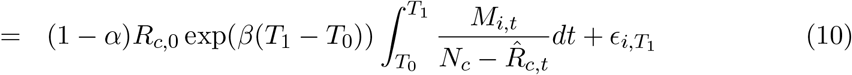

This equation allows us to solve for (1−*α*) exp(*β*(*T*_1_−*T*_0_) if we observe *R*_*c*,0_. We will allow for observing *R*_*c*,0_ with error in Section 3.3. We can estimate *β* from the growth of reported infections in the epicenter because there is no influx of infected people from other regions. Given that *β* is now determined, we can solve for *α*. However, there is much variation in estimation of *β* within the literature, and our estimate of *α* varies with point estimates of *β* (Liu et al., 2020; Read et al., 2020; Shen et al., 2020).

### 3.3 When travel data and a random testing benchmark are available

In this scenario, we will be leveraging the same fact that the number of incoming unreported infections is informed by the travelers from the epicenter. Now we can also allow for selection in traveling. More specifically, if we think that e.g. urban areas are likely to have a higher infection rate than rural areas and travel abroad more^11^, then assumption 3.1 might not hold. Therefore, we introduce a bias correction term *γ* in the relation between the fraction of infected among travelers and the fraction of unreported infected individuals in the general population. This bias correction term *γ* can also account for the fact that a fraction of the unreported infected people might be too sick to travel. Our relaxed assumption in this scenario is:

#### Assumption 3.2.

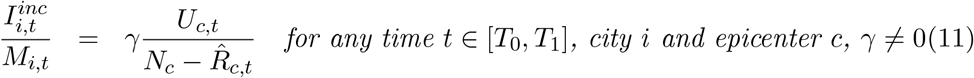

We can further allow for the fact that the reporting rate in epicenter *α*_*c*_ can be different from that of region *i*, so 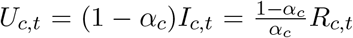. We can now rewrite assumption 3.2 as:

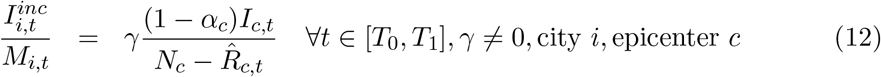

Plugging into equation (1) and (3), we get

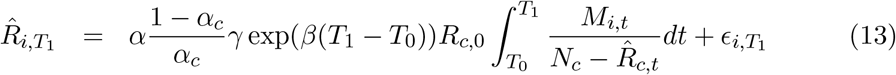

The additional parameters for the bias correction term *γ* and different reporting rate for the epicenter complicate the estimation of *α* using travel data alone. However, having data from a country that has done randomized or complete testing greatly helps overcome this challenge. In our case, we are able to identify *α* using additional information given by the randomized testing benchmark provided by Iceland. Since the Iceland company deCODE genetics implemented random testing of COVID-19 for a representative sample of the island population^12^, we are able to observe true infection rate of Iceland at time *T*_1_. Multiplying this true infection rate by the population of region *j* in Iceland will give us the actual number of infections in region *j* at time *T*_1_, which is 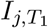. Thus, for any region *j* in Iceland we observe 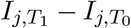, which in turn, equals the infections generated by travelers from the epicenter:

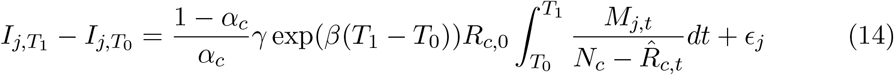

Note that if we allow for iid measurement error *ϵ*_*j*_ in observing 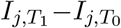, we can get a consistent estimate of 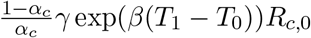 by estimating equation (14). If we don’t allow for measurement error *ϵ*_*j*_, then we can estimate 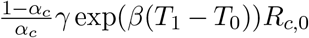 with no error. Estimating equation (13) gives consistent estimate of 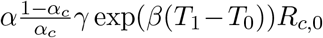. Taking the ratio, we have identified *α*.

One intuition for this strategy is the following: the ratio between travel to U.S. and travel to Iceland from the epicenter should tell us the ratio of total infections between U.S. and Iceland. Iceland’s randomized testing gives us its number of total infections, so U.S. total infections can be computed. In other words, we observe the outcome in U.S. with under-reporting, and the unobserved counterfactual outcome with full reporting is given by the benchmark Iceland. An additional advantage of this estimation/identification strategy, as opposed to the previous strategy in section 3.2, is that now we don’t need an estimate of *β* in order to recover *α*. We also allow for the fact that *R*_*c*,0_ could be observed with error. Identifying *α* does not require observing *R*_*c*,0_ perfectly because *R*_*c*,0_ appears identically in both equations.

We should be clear that for terms with *β* to cancel out, the argument does assume that *β* is the same across Iceland and the U.S. We believe this might be a reasonable assumption for the early periods of the infection when social distancing or other widespread measures had not yet been implemented (in a potentially differential fashion). We also need the bias term *γ* to be the same for US and Iceland; this means that proportion of (unreported) infected travelers from China to the U.S. and Iceland are the same. More detailed micro-data on travelers may be used to assess the validity of this assumption.

This estimation strategy also works when a complete testing benchmark exists. If the whole population of region *j* is tested, then we observe 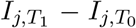 trivially. Equation (14) still gives consistent estimate of 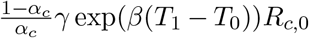 and the rest of the argument follows.

Note that if in the model reported infections and unreported infections have different transmission rates, then our strategy would not be able to capture the differential rates.

We would need other sources of information to help us pin down these differential rates.

### 3.4 Incorporating Reporting Lags

In this section, we show how our model can incorporate a fixed reporting lag in reported infections and derive identification equations. Reporting lags are important, because if people are tested for the virus only after symptoms show up, there will be a lag in reported infections. Another major reason for reporting lag is the lag in testing results. The turnaround time for testing results in U.S. major laboratory companies could be 2 to 3 days (Kaplan and Thomas, 2020).

We denote true infected, true reported infected, and true unreported infected in time *t* and target city *i* as *I*_*i,t*_, *R*_*i,t*_, *U*_*i,t*_ respectively. Those for epicenter *c* as *I*_*c,t*_, *R*_*c,t*_, *U*_*c,t*_. Let *k* be the lagged report period. At time *t* city *i* denote the lagged reported infected *LR*_*i,t*_ = *R*_*i,t*−*k*_. For epicenter *c*, lagged reported infected is *LR*_*c,t*_ = *R*_*c,t*−*k*_. But the observed lagged reported infected of target city 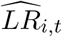 is with iid measurement error *ϵ*_*i,t*_.

Define reporting rate at city *i* as 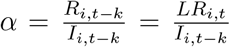 and at epicenter *c* as 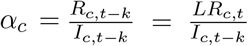. This means that we are considering the reporting rate of lagged reported cases as a fraction of the lagged total infections.

When travel data are available but randomized testing data unavailable, we still maintain assumption 3.1. In city *i* time *T*_1_, the estimating equation is

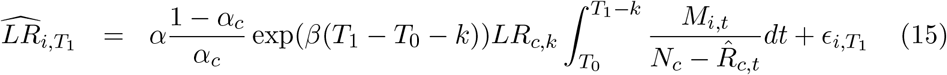

If *α* = *α*_*c*_, then both of them are identified if *β* and *k* are identified and *LR*_*c,k*_ is observed without error.

When both travel data and randomized testing data are available, we maintain assumption 3.2. In US city *i* time *T*_1_, the estimating equation is

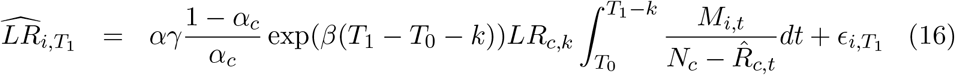

In Iceland region *j* time *T*_1_, the estimating equation is

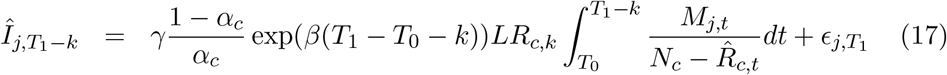

If we know *k*, then we can compute 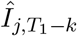 from the randomized testing data. Regressing equation (17) gives consistent estimate of 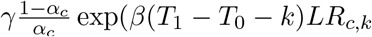. Regressing equation (16) gives a consistent estimate of 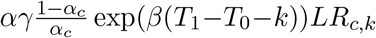. Taking the ratio, we can identify *α* if we know *k*. Again here we can also allow for the situation where we don’t observe *LR*_*c,k*_ perfectly. Details of how we derive the estimating equations are in the appendix.

## 4 Data

### 4.1 COVID-19 Data

Daily reported infections and recovery data are collected by Johns Hopkins University of Medicine Coronavirus Resource Center from January 22 to March 31, 2020. We use data for all U.S. states/counties and Iceland.

Randomized testing data in Iceland is obtained from the website maintained by the Directorate of Health and the Department of Civil Protection and Emergency Management in Iceland^13^. We have daily number of tests conducted by deCODE genetics and daily number of confirmed cases. We use the first wave of randomized testing by deCODE which spans March 15 − 19, 2020. During the first wave they performed 5490 tests and confirmed 48 cases, which implies an infection rate of .874%. The randomized testing conducted was random over non-confirmed individuals which made up a very tiny portion of the Icelandic population at this time. This could lead to slightly downward biases in our Iceland confirmed data, slightly biasing our US alpha estimates upward.

In our main estimation we consider February 23 as *T*_0_ and March 10 as *T*_1_ with a 5 day lag, and *T*_1_ as Mar 13 for an 8 day lag. This is because there were very few infections in January and early February. We check robustness of different time periods in Section 5.3.

### 4.2 Travel Data

We obtain monthly data of international arrivals to U.S. by port of entry and country of origin from I-94 Arrivals by National Travel and Tourism Office. We use the number of visitors from China, Italy, Spain, UK, and Germany in January and February 2020 as the measure for incoming travelers to U.S. states. For international arrivals to Iceland, we get the number of visitors from China, Italy, Spain, UK, and Germany in January and February 2020 from the Icelandic Tourist Board. We have not been able to obtain March travel data into either country.

The National Travel and tourism office of the United States provides monthly data for entry by port of entry, as well as a separate data set for country of origin. We construct the number of visitors from China, Italy, Spain, Germany, and the UK by scaling the port-of-entry data by the percentage of total visitors that are from these countries. This introduces error, as we cannot observe directly the number of e.g. Chinese travelers into a particular city or state. It is also important to note that we do not observe inter-state travel. While this may not be important for the immediate infections caused by travelers from the epicenter, our projections for the number of infections for *T*_1_ that are far removed from *T*_0_ will be less accurate due to interstate travel.

For Icelandic data, 99% of international travelers arrive through Keflavik airport into Iceland. The data contains a breakdown of arrival by country of origin, broken down by month of arrival. We use January and February arrival data from China, Italy, Spain, UK, and Germany for estimation.

Our travel data for both countries does not control for connecting flights. However the United States data is limited to the top-30 port of entries, many of which are large urban cities for which there will be less connecting flights. Further work that can obtain more precise estimates of entry may be able to control for this. In the case of Iceland: a survey conducted by the Icelandic Tourist Board suggests that 2-5% of international travelers are aboard connecting flights, suggesting it is less of a problem for this set of data.

### 4.3 Population Data

Estimates of U.S. State and county population data come from the U.S. Census Bureau. Data for the populations of China, Iceland, Italy, Spain, UK, and Germany as of 2020 are obtained from the United Nations Population Division.

## 5 Empirical Application

### 5.1 Implementation

We now consider estimation of *α*_*US*_ using randomized sampling in Iceland, as described in Section 3.3. Randomized sampling done by deCODE genetics gives a percentage of the population that has contracted the virus. We estimate equation (14) using Randomized Testing to construct 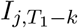. We do not have city-level travel data into Iceland. 99% of all international travel arrives through a single airport, and while the data provided is accurate, this gives only a single data point for estimation. As a result, equation (14) relies on a single data point of travel and infection, but 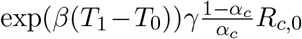 is estimated without error by the ratio of 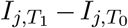 and 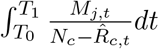.

For estimation of reporting rates in the U.S., we need estimates of several figures: Firstly *M*_*j,t*_, and secondly of 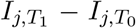. We discuss the estimation of these here. We observe only monthly travel data to construct *M*_*j,t*_, and to maintain robustness to January travels and infections, we average February and January travel into both the United States and Iceland. We assume that *M*_*j,t*_ is uniform over the entire time period such that 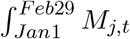 is equal to the sum of all travel into the city from January and February. Thus the integral 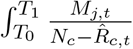 varies only due to the confirmed infections increasing over time. Estimation of 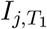 is complicated due to randomized testing by Iceland only being conducted at certain dates. To resolve this problem, we scale the Iceland randomized results by the scale of the confirmed cases against March 15. This means that if there were half the confirmed cases in March 5 as in March 15, the total infections would be half of the randomized testing percent times the population of Iceland. This allows for us to consider *T*_1_ closer to the onset of the infection than the randomized testing dates. We also remove the number of infected from Wuhan China from our data on confirmed infected in China due to the lock-down restrictions placed on this city. We use the first wave of deCODE testing to determine the percentage of the population that has contracted the disease. This testing took place during Mar 15 through Mar 19. The results show that .874% of the population of Iceland have contracted the disease as of Mar 15^14^.

We estimate equation (13) using multiple data points from U.S. states and counties. We obtain our estimate of 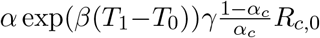 via OLS without a constant term. One important note is that if the magnitude of measurement error in travel data were high, this problem may be alleviated via instrumental variables strategy using other travel data measured with error.

We then construct our estimate of *α* by dividing the two estimates. It is important to note that as a result of the division, this method is not reliant on population data from the epicenter of infection. As long as *γ* and *β* are the same between Iceland and the United States we will have identified *α*. It is likely that at the onset of the infection similar preventative measures have been taken in these two countries, meaning that *β* will be reasonably close for each country.

Is China the only epicenter for the United States? While the first confirmed infection in Seattle occurred from a visitor from China, our data on The United States and Iceland occurs later in the global progression of the virus than our Chinese data.

By the time these countries were experiencing infections, Italy had also experienced an outbreak. To this end, we also allow for a second epicenter: Italy. Italy is located much closer to Iceland and constitutes a substantial amount of travel to the country. However, to maintain identification, we require that *α, β* and *T*_0_ be same for both China and Italy, and we observe *LR*_*c,k*_ for both epicenters with no error. However we find that allowing *T*_0_ to vary does not affect our estimates by much. We also consider a broader collection of epicenters of China, Italy, Spain, Germany and the UK. For some collection of epicenters *L*: Our estimation equation for the United States is given below.

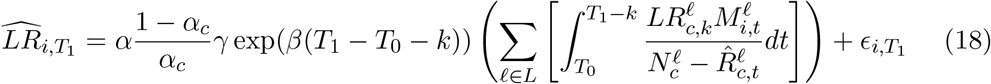

A similar equation is also estimated with multiple epicenters (China and Italy, also with Spain, Germany, and UK) for Iceland.

### 5.2 Results: Illustration

As a first illustration of our approach, we first estimate *α* using February 23 as *T*_0_ and March 10 as *T*_1_. We consider other dates for robustness later. We consider two lag models, 5 days, the median time for symptoms to appear, and 8 days, to capture the testing lag in addition to symptom onset (Lauer et al. (2020); Kaplan and Thomas (2020); Li et al. (2020)). For the 8 day lag, *T*_1_ was set to March 13 for comparison. We estimate, for the 8 day lag, a value of *α* = 0.0416 (s.e. 0.00984).^15^ This would mean that for every case confirmed in the United States in early March, there are still 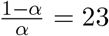 unconfirmed cases (assuming a reporting lag of 8 days).

There is one city present in our data that is a huge outlier. Seattle featured very early infections, and was unable to contain the spread of early infections unlike other cities in the United States. We believe that for King County, *T*_0_ may be much earlier than for the other cities. This means that within our time interval, there are substantial amounts of infections caused by residents of the city, not only visitors. As a result, this city has a substantially higher (3700%) amount of confirmed cases per visitor than any other city at the current time, and we exclude it from the data.

Correct estimation of the reporting lag parameter is essential, as our estimates of *α* are sensitive to this. We consider its robustness in the following section.

Our approach is also sensitive to the travel data magnitudes, which may not be well estimated for the United States due to data limitations. In particular, connecting flights after port of entry may lead to underestimates of international arrivals into smaller cities and counties. We also lack inter-state travel between the United States, which would be important for estimating *α* later into the spread of the virus.

Have we considered all epicenters of the virus for the United States and Iceland? There were other countries which had seen substantial infections such as South Korea. Their exclusion biases both the estimates from both Iceland as well as the United States, and as long as the magnitudes of travel were even between the two will not bias alpha. If these other epicenters had more travel to the United States relative to Iceland United States than Iceland, this would downward bias our estimates of *α*, and vice versa. However, we see little change in our estimates by adding in Spain, Germany and UK. If the travel patterns between the United States and Iceland to and from an omitted set of epicenter countries are not very different, we do not believe their omission will substantially alter our results.

### 5.3 Results: Range of Estimates and Robustness Checks

Our dates for *T*_0_ and *T*_1_ are chosen such that they capture the onset of the infection for the United States. As table 3 shows, our *α* estimate is reasonably stable along choices of *T*_1_, and very stable among choices of *T*_0_ all throughout February. We estimate a range of 1.5% − 10% for the average reporting rate across the US with a reporting lag of 5 days and 4% − 14% reporting rates when there is a lag of 8 days. Using only China as the epicenter, we observe similar patterns in *α*. For early March we note a relatively stable *α* over *T*_1_. For very early choices for *T*_1_, our Iceland estimates of confirmed are very small, and this could create very noisy estimates of *α* (the first case in Iceland was confirmed February 28). As we increase *T*_1_, we see an increase in *α*. This may be due to increases in the availability of test kits, which lead to higher reporting rates. However, this result may in part be due to unobserved/unaccounted travel, particularly within the United States, along with the fact that we do not have data on March travel into the US. Both of these factors would lead to under-reporting of travel for late March, and cause estimates of *α* to be upward biased. Moreover, as we progress later into March, social distancing/health policy measures across Iceland and U.S. began to be applied, leading to differential changes in the transmission rate.

How would these estimates affect our estimate of total infections, as opposed to reported infections? As an illustration, with our estimated average reporting rates from table 3, we compute the estimated total infections for different U.S. counties as of March 15 and March 20 in table 5. To give a range of our estimates, we take the 10th and 90th percentile of the estimated reporting rates over all cutoffs to compute a bound on estimated total infections. As of March 20, our results suggest, for our average *α* estimate in table 3, that there were 41,205 (85,937) Infected residents of New York City, with a 10th percentile of 24,366 (47,257) and a 90th percentile of 114,849 (306,607) for an assumed 8 (5) day lag. These estimates imply that, as of March 20, with an assumed 8(5) day reporting lag, about 0.5% (1%) of New York City population was infected for our average estimate in table 2, and 1.4% (3.6%) at the 90th percentile of our estimates.

**Table 5:** Estimated Total Infected By County - Lag 5

| County | Mar 15 |  |  |  | Mar 20 |  |  |  |
| --- | --- | --- | --- | --- | --- | --- | --- | --- |
|  | Rep. | 10Pct. | Mean | 90Pct. | Rep | 10Pct. | Mean | 90Pct. |
| Broward, FL | 36 | 330 | 601 | 2143 | 128 | 1174 | 2135 | 7619 |
| Clark, NV | 16 | 147 | 267 | 952 | 126 | 1156 | 2102 | 7500 |
| Cook, IL | 50 | 459 | 834 | 2976 | 278 | 2550 | 4638 | 16548 |
| Fulton, GA | 20 | 183 | 334 | 1190 | 88 | 807 | 1468 | 5238 |
| Harris, TX | 11 | 101 | 184 | 655 | 30 | 275 | 501 | 1786 |
| Hillsborough, FL | 4 | 37 | 67 | 238 | 32 | 294 | 534 | 1905 |
| Honolulu, HI | 3 | 28 | 50 | 179 | 28 | 257 | 467 | 1667 |
| Los Angeles, CA | 53 | 486 | 884 | 3155 | 292 | 2679 | 4872 | 17381 |
| Maricopa AZ | 4 | 37 | 67 | 238 | 34 | 312 | 567 | 2024 |
| New York City, NY | 269 | 2468 | 4488 | 16012 | 5151 | 47257 | 85937 | 306607 |
| Philadelphia, PA | 4 | 37 | 67 | 238 | 67 | 615 | 1118 | 3988 |
| Ramsey, MN | 5 | 46 | 83 | 298 | 16 | 147 | 267 | 952 |
| San Diego, CA | 16 | 147 | 267 | 952 | 127 | 1165 | 2119 | 7560 |
| San Francisco, CA | 28 | 257 | 467 | 1667 | 76 | 697 | 1268 | 4524 |
| Suffolk, MA | 27 | 248 | 450 | 1607 | 86 | 789 | 1435 | 5119 |

Throughout the analysis above, we have excluded King County, Washington which contains Seattle. Table 7 displays our estimates including this county, which heavily skews the data. We believe this may be due to significant community infections occurring in the county during our time period, as the city was infected much earlier than other cities.

**Table 6:** Estimated Total Infected By County - Lag 8

| County | Mar 15 |  |  |  | Mar 20 |  |  |  |
| --- | --- | --- | --- | --- | --- | --- | --- | --- |
|  | Rep | 10Pct. | Mean | 90Pct. | Rep | 10Pct. | Mean | 90Pct. |
| Broward, FL | 36 | 170 | 288 | 803 | 128 | 605 | 1024 | 2854 |
| Clark, NV | 16 | 76 | 128 | 357 | 126 | 596 | 1008 | 2809 |
| Cook, IL | 50 | 237 | 400 | 1115 | 278 | 1315 | 2224 | 6198 |
| Dallas, TX | 11 | 52 | 88 | 245 | 74 | 350 | 592 | 1650 |
| Essex, NJ | 7 | 33 | 56 | 156 | 73 | 345 | 584 | 1628 |
| Fulton, GA | 20 | 95 | 160 | 446 | 88 | 416 | 704 | 1962 |
| Harris, TX | 11 | 52 | 88 | 245 | 30 | 142 | 240 | 669 |
| Hillsborough, FL | 4 | 19 | 32 | 89 | 32 | 151 | 256 | 713 |
| Honolulu, HI | 3 | 14 | 24 | 67 | 28 | 132 | 224 | 624 |
| Los Angeles, CA | 53 | 251 | 424 | 1182 | 292 | 1381 | 2336 | 6511 |
| Maricopa AZ | 4 | 19 | 32 | 89 | 34 | 161 | 272 | 758 |
| Miami-Dade, FL | 13 | 61 | 104 | 290 | 123 | 579 | 980 | 2731 |
| Multnomah, OR | 1 | 5 | 8 | 22 | 12 | 57 | 96 | 268 |
| New York City, NY | 269 | 1272 | 2152 | 5998 | 5151 | 24366 | 41205 | 114849 |
| Philadelphia, PA | 4 | 19 | 32 | 89 | 67 | 317 | 536 | 1494 |
| Ramsey, MN | 5 | 24 | 40 | 111 | 16 | 76 | 128 | 357 |
| San Diego, CA | 16 | 76 | 128 | 357 | 127 | 601 | 1016 | 2832 |
| San Francisco, CA | 28 | 132 | 224 | 624 | 76 | 360 | 608 | 1695 |
| Suffolk, MA | 27 | 128 | 216 | 602 | 86 | 407 | 688 | 1918 |
| Wayne, MI | 8 | 38 | 64 | 178 | 67 | 317 | 536 | 1494 |
| Whatcom, WA | 2 | 9 | 16 | 45 | 10 | 47 | 80 | 223 |

**Table 7:** Reporting Rate (*α*) Estimates Including King County

| | $\alpha_{ice}$ | $\alpha_{US}$ | $\frac{1-\alpha_{US}}{\alpha_{US}}$ |
| --- | --- | --- | --- |
|  | 5 Day Lag |  |  |
| China and Italy Travel Data | 0.0231 | 0.0206 | 47.6 |
| Only Chinese Travel Data | 0.0231 | 0.0211 | 46.5 |
| China, Italy, Spain, Germany, UK | 0.0231 | 0.0211 | 46.5 |
|  | 8 Day Lag |  |  |
| China and Italy Travel Data | 0.0231 | 0.0516 | 18.4 |
| Only Chinese Travel Data | 0.0231 | 0.0539 | 17.6 |
| China, Italy, Spain, Germany, UK | 0.0231 | 0.0538 | 17.6 |

We report estimated $\alpha$ by OLS without a constant for several specifications of the model. We use $T_0$ as Feb 23rd and $T_1$ as Mar 10,13 for each lag respectively. For the versions including European data, European travel to both Iceland and the United States is considered. King County is included in this data.

We consider our estimates robustness to reporting lags in table 8. Our estimates of *α* appear reasonably robust to a range of lengths of the lag, with an increase as the lag becomes longer.

**Table 8:** Robustness to Lag

| Lag | $\alpha$ |
| --- | --- |
| 0 | 0.00558 |
| 1 | 0.00874 |
| 2 | 0.0094 |
| 3 | 0.01 |
| 4 | 0.0123 |
| 5 | 0.0168 |
| 6 | 0.0287 |
| 7 | 0.0299 |
| 8 | 0.0452 |
| 9 | 0.0717 |
| 10 | 0.0758 |
| 11 | 0.124 |
| 12 | 0.211 |

Table 8 shows estimates of $\alpha$ as the reporting lag period is varied. We use $T_0$ as Feb 23, and $T_1$ as March 5 + Lag days. King County is omitted. We include Italy, Spain, Germany, and the United Kingdom as epicenters as well as China.s

We note that large lags (*k >* 10) pose a problem for estimation in our model. For estimation purposes, we maintain *T*_1_ − *k* to be a constant date as we consider changes in the lag parameter. This means that for large lags, we must consider *T*_1_ dates deep into March. However, the further we get into March, the more interstate travel and carrying of infections between cities and states matters, which may lead to overstating *α*. To complicate matters, Icelandic and the US policies for handling the spread of the virus may have diverged significantly. This means that our assumption of *β* constant between countries also may not hold. When it does not, our estimate of *α* identifies *α* × exp [(*β*_*US*_ − *β*_*I*_) (*T*_1_ − *T*_0_)]. *β*_*US*_ *> β*_*I*_ implies that we are overestimating *α* for large *T*_1_ values. Evidence from Kucharski et al. (2020) suggests that *β* is very sensitive to changes in policy, leading to this upward bias in *α*.

## 6 Conclusion

In this paper, we lay out a simple model of disease transmission across a known epicenter and target cities. Using this model, we provide simple analytical arguments to allow the estimation/identification of reporting rates in target cities away from the epicenter. Our preferred estimation strategy utilizes variation of travel patterns from epicenter to destination cities and available randomized testing results from elsewhere in the world. The empirical implementation of our model generates a range of estimates for the percentage of infections that have been reported. Using international travel data to the U.S. and randomized testing data from Iceland, for the February to early March window, our estimates of the average reporting rate in the U.S. lie in a range of 4 − 14%, accounting for an assumed reporting lag of 8 days. (The range is 1.5 − 10%, accounting for a reporting lag of 5 days.) Our estimates suggest that a large number of infections in the U.S. have not been reported in this early period.

We should be very clear that we are not offering or endorsing any policy recommendations based on our estimates. Nor do we suggest that any of our analysis should be taken as a substitute for randomized/complete testing, which will provide the most reliable estimates of the true infection rate in the population. Our primary aim in this paper has been to obtain tractable analytic results showing how to identify the reporting rate from available data. We also note that our model is a substantially stripped down version of epidemiological models considered by (Li et al., 2020; Wu et al., 2020; Flaxman et al., 2020). These more complex models may allow additional sources of variation in the data to pin down the key parameters of interest. Importantly, we do want to emphasize that our identification and estimation results rely quite sensitively on model assumptions and the (un)availability of high quality data on travel. We hope future research can improve on these important limitations.

**Figure 1:**
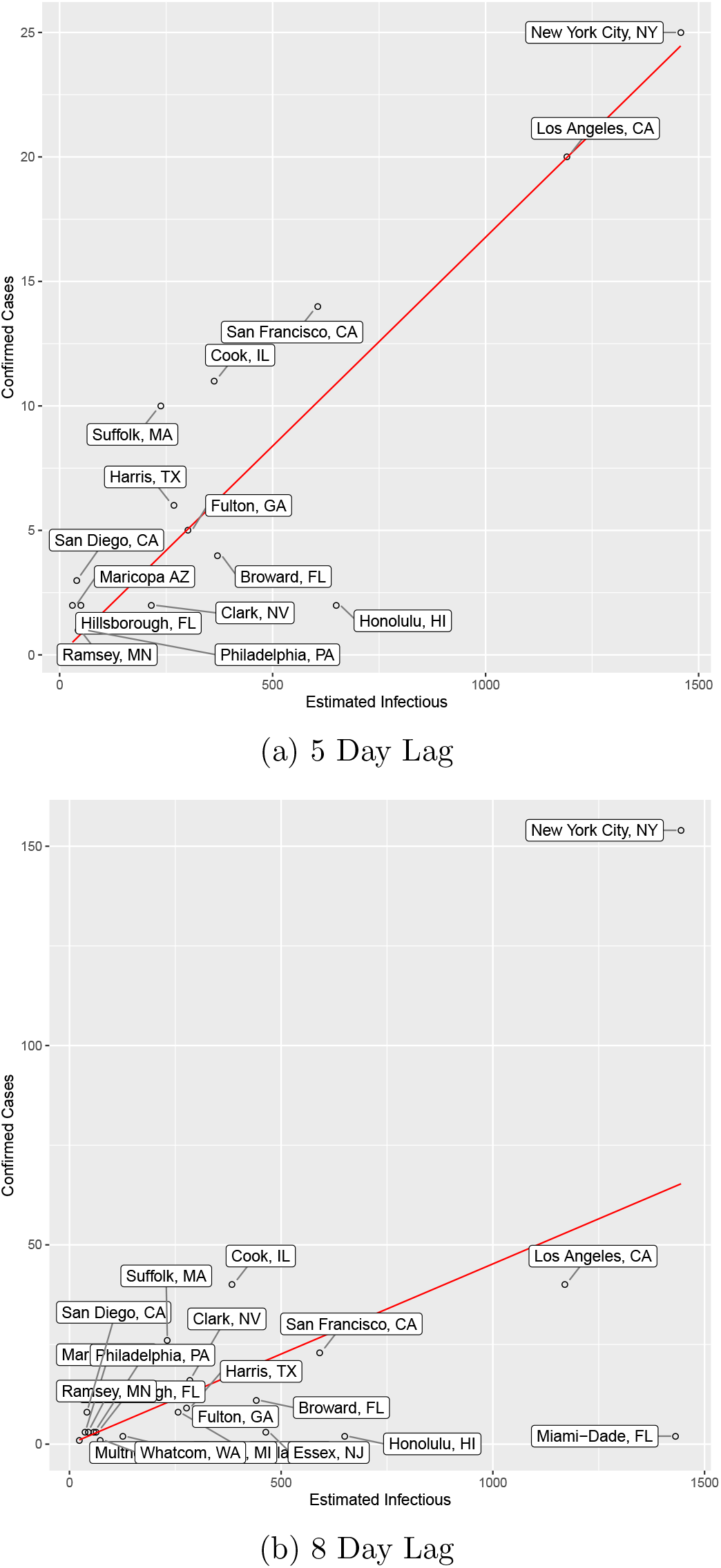
Estimated reported infections by county. This plot shows the ratio of Confirmed Cases to Estimated Cases in each County on *T*_1_ = March 10 for 5 day lag, and 13 for 8 day lag. *T*_0_ is February 23rd. We use the full European Entry. Estimated Cases are computed as the ratio of the right hand side of equation 16 and the ratio of equation 17

## Data Availability

Data and code for replication are available at the link below.

https://github.com/tschwieg/Covid19-ReportingEstimation

## Appendix

We derive our model incorporating reporting lags in the appendix and show how we get the estimating equations in Section 3.4.

Recall that we denote true infected, true reported Infected, and true unreported infected in time *t* and target city *i* as *I*_*i,t*_, *R*_*i,t*_, *U*_*i,t*_ respectively. Those for epicenter *c* as *I*_*c,t*_, *R*_*c,t*_, *U*_*c,t*_. Let *k* be the lagged report period. At time *t* city *i* denote the lagged reported infected *LR*_*i,t*_ = *R*_*i,t*−*k*_. For epicenter *c*, the lagged reported infected is *LR*_*c,t*_ = *R*_*c,t*−*k*_.

Define reporting rate at city *i* as 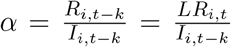 and at epicenter *c* as 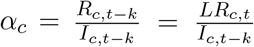. This means that we are considering the reporting rate of lagged reported cases on the lagged total infection.

We know that in the epicenter *c*, we have the following:

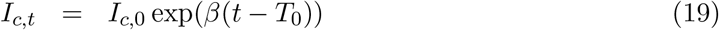

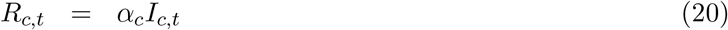

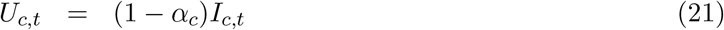

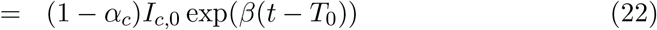

When only travel data is available, our assumption 3.1 is

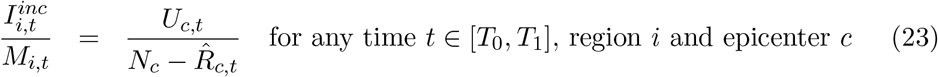

We can then write it as

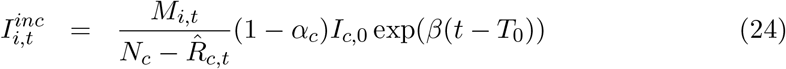

In city *i*, at time *T*_1_ we observe 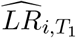. We have

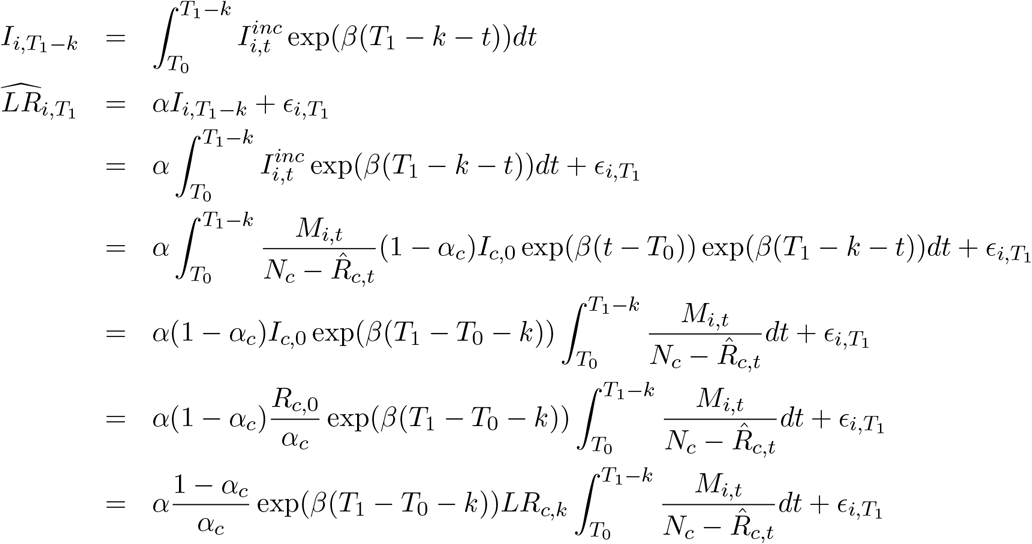

When both travel data and randomized testing data are available, we maintain assumption 3.2:

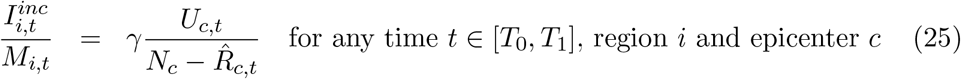

We can write it as

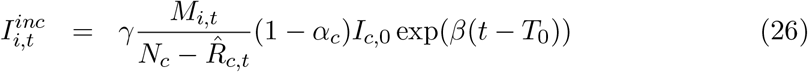

In US city *i*, at time *T*_1_ we observe 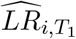. Following the same derivation as above, we have

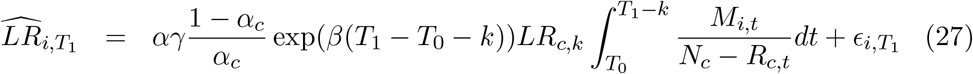

Similar derivation shows that for Iceland region *j* time *T*_1_, we have

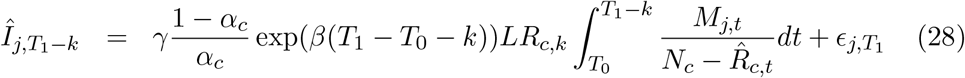

## Footnotes

1 https://www.whitehouse.gov/presidential-actions/proclamation-declaring-national-emergency-concerning-novel-coronavirus-disease-covid-19-outbreak/

2 https://www.wsj.com/articles/a-state-by-state-guide-to-coronavirus-lockdowns-11584749351

3 https://www.ft.com/content/01f267a2-686c-11ea-a3c9-1fe6fedcca75

4 https://www.who.int/docs/default-source/coronaviruse/situation-reports/20200306-sitrep-46-covid-19.pdf?sfvrsn=96b04adf_2

5 Of course, another strategy is to assume that the reporting rate discovered through randomized testing in Iceland is the same in the destination city/country of interest (Section 3.1).

6 This assumes that cases are reported with a lag of 8 days as in Table 3(b), and incorporates travel data from China, Italy, Spain, Germany, and the UK. A shorter assumed reporting lag of e.g. 5 days generates a range of estimated reporting rates between 1.5% to 10%. We have excluded King county, Washington in these results because this county containing Seattle shows much earlier community infections than other regions in U.S.

7 For example, that Li et al. (2020) estimates different transmission rates for reported vs. unreported infections, which we are unable to identify with our strategy. Li et al. (2020) assume that unreported infected individuals transmit the disease at a slower rate than reported infected individuals. However, since most reported infections are either hospitalized or self-quarantined, it is not clear whether this assumption is an *a priori* reasonable one.

8 Bogoch et al. (2020) and Lai et al. (2020) calculates how vulnerable countries are to the virus by the magnitude of travelers from Wuhan, and correlate these vulnerability/risk measures with reported cases in these countries.

9 This measurement error *ϵ*_*i,t*_ could arise from error in collecting the data.

10 In reality *α* may be varying over time, due to e.g. changes in the extensiveness of testing. If this is the case, as we vary the [*T*_0_, *T*_1_] window, we will obtain window-specific estimates of *α*, which can be thought of as a weighted average of *α* during this period.

11 This is likely to be true in epicenters like China.

12 We will describe the randomized testing in detail in Section 4

13 https://www.covid.is/data

14 Stock et al. (2020) also estimates the undetected rate and total infection rate in the Iceland study.

15 We are reporting a naive OLS standard error here. We have not fully explored the sampling properties of this estimation method.

